# Sputum microbiome α-diversity is a key feature of the COPD frequent exacerbator phenotype

**DOI:** 10.1101/2023.08.09.23293835

**Authors:** Alexa A. Pragman, Shane W. Hodgson, Tianhua Wu, Allison Zank, Cavan S. Reilly, Chris H. Wendt

## Abstract

**Background:** The lung microbiome is an inflammatory stimulus whose role in chronic obstructive pulmonary disease (COPD) pathogenesis is incompletely understood. We hypothesized that the frequent exacerbator phenotype is associated with decreased α-diversity and increased lung inflammation. Our objective was to assess correlations between the frequent exacerbator phenotype, the microbiome, and inflammation longitudinally during exacerbation-free periods.

**Methods:** We conducted a case-control longitudinal observational study of the frequent exacerbator phenotype and characteristics of the airway microbiome. Eighty-one subjects (41 frequent and 40 infrequent exacerbators) provided nasal, oral, and sputum microbiome samples at two visits over 2-4 months. Exacerbation phenotype, relevant clinical factors, and sputum cytokine values were associated with microbiome findings.

**Results:** The frequent exacerbator phenotype was associated with lower sputum microbiome α-diversity (p=0.0031). This decrease in α-diversity among frequent exacerbators was enhanced when the sputum bacterial culture was positive (p<0.001). Older age was associated with decreased sputum microbiome α-diversity (p=0.0030). Between-visit β-diversity was increased among frequent exacerbators and those who experienced a COPD exacerbation between visits (p=0.025, p=0.014). Sputum cytokine values did not differ based on exacerbation phenotype or other clinical characteristics. IL-17A was negatively associated with α-diversity, while IL-6 and IL-8 were positively associated with α-diversity (p=0.012, p=0.012, p=0.0496). IL-22, IL-17A, and IL-5 levels were positively associated with *Moraxella* abundance (p=0.027, p=0.0014, p=0.0020).

**Conclusions:** Even during exacerbation-free intervals, the COPD frequent exacerbator phenotype is associated with decreased sputum microbiome α-diversity and increased β-diversity. Decreased sputum microbiome α-diversity and *Moraxella* abundance are associated with lung inflammation.

## INTRODUCTION

Chronic obstructive pulmonary disease (COPD) is a leading cause of death, however the mechanisms driving its progression remain incompletely understood. One recently recognized mechanism is inflammation triggered by the lung microbiome. Cycles of recurrent lung infection, inflammation, and antibiotic use may disrupt the microbiome with downstream consequences for lung function [1]. Even in the absence of clinical infection, COPD patients with pathogenic bacteria in their airways have higher levels of sputum and systemic inflammatory markers and increased pulmonary symptoms [2, 3].

The COPD frequent exacerbator phenotype identifies a subset of patients at high risk of recurrent COPD exacerbation. Frequent exacerbators (FE) suffer increased morbidity and mortality compared to those who experience exacerbations less often [4–7]. Approximately half of COPD exacerbations are attributed to bacterial infection, and pathogenic bacteria such as *Haemophilus influenzae, Moraxella catarrhalis*, and *Streptococcus pneumoniae* are often identified in the lung microbiome of COPD patients even during periods of stable lung disease [3].

Inflammatory markers are increased in the sputum of FE [8–10] and COPD patients colonized with potentially pathogenic bacteria [9, 11–15]. The COPD lung microbiome provides an inflammatory stimulus, even in the absence of overt lung infection. In particular, IL-17A, IL-8, IL-6, IL-1β, IL-22, IL-5, and leukotriene B4 (LTB4) levels in sputum have been associated with various components of the COPD lung microbiome during exacerbation-free intervals [2, 8, 11–19]. It remains unclear which particular components of the microbiome (bacterial biomass, α-diversity, the particular taxa present, etc.) are most closely associated with sputum inflammation or the FE phenotype.

We and others have shown that the sputum microbiome of FE has lower alpha diversity compared to non-FE [18, 20–26]. Few of these studies, however, evaluate the COPD lung microbiome solely during periods of clinical stability (when findings are less influenced by exacerbation treatments), longitudinally, and including an analysis of concurrent lung inflammation. We undertook the present case-control longitudinal observational study of COPD exacerbation phenotype, the upper airway and sputum microbiome, and lung inflammation to address these gaps.

## METHODS

### Study design and recruitment

We conducted a case-control longitudinal observational study of exacerbation phenotype and characteristics of the upper airway and sputum microbiome. All participants were recruited from a single site and were age 40 or older with COPD. Frequent exacerbators (FE) had at least one severe exacerbation (an exacerbation requiring hospital admission or emergency department visit) in the last 12 months, in accordance with descriptions found in GOLD guidelines [27]. Infrequent exacerbators (IE) must have had 0 exacerbations in the prior 24 months. Recruitment and all visits/samples were deferred until participants had recovered for at least 1 month from the most recent exacerbation. The protocol was approved by the Minneapolis VA IRB (#4541-B). Additional details on recruitment and other methods can be found in the supplementary information. Samples from 22 participants have been previously published (PRJNA543785) [20], however all data analyzed here were re-sequenced for this analysis (PRJNA944199).

### Study procedures

At visit 1, participants provided their medical history, underwent spirometry, completed the St. George’s Respiratory Questionnaire (SGRQ), and provided oral wash, nasal swab, and induced sputum samples. All participants returned for a second study visit ∼2 months following visit 1, where they provided information on interim COPD exacerbations, repeated the SGRQ, and provided oral, nasal, and sputum samples. Visit 2 was deferred (for up to 4 months following visit 1) if the participant reported any COPD exacerbations or antibiotic use in the 1 month prior to the visit. Exacerbation phenotype was determined at visit 1 and was not revised based on exacerbations observed during the study.

### Sample processing, 16S rRNA gene quantification, and MiSeq sequencing

All samples and negative controls were extracted using the MO BIO PowerSoil DNA Isolation Kit (QIAGEN, Germantown, MD). Extracted DNA underwent 16S rRNA gene quantification using droplet digital PCR (ddPCR) and 16S rRNA gene V4 MiSeq sequencing. 16S rRNA V4 sequences were processed as described in the supplementary information.

### Sputum culture results

The clinical microbiology laboratory performed Gram stain and aerobic culture on all sputum samples. Any organism identified in culture was considered a pathogen.

### Cytokine analyses

Sputum samples were submitted to the University of Minnesota Cytokine Reference Laboratory for determination of LTB4, GM-CSF, IL-8, IL-6, IL-1β, IL-17A, IL-22, IL-5, and TNF-α (R&D Systems, Minneapolis, MN USA).

### Statistical analyses

Data presented is from visit 1 only (V1) or both visits (BV), as described in the text. Linear regression (LR), including random effects censored regression models, linear mixed models (LMM), and generalized estimating equations (GEE), were used to test for associations between variables. Analyses of α-diversity metrics were adjusted for age, forced expiratory volume in 1 second percent predicted (FEV1pp), BMI, current tobacco use, and current alcohol use. β-diversity was assessed via the Bray-Curtis dissimilarity metric and illustrated via principal coordinates analysis (PCoA). Univariate PERMANOVA analyses of β-diversity were performed for each anatomical site. Permutation tests were used to determine if within-subject visit 1-visit 2 similarity differed by permuting levels of categorical clinical factors. After eliminating genera present in <10% of samples, tests of association between taxa and clinical characteristics used Holm’s procedure to control the family-wise error rate across all taxa at 5%. GM-CSF and TNF-α were not analyzed and IL-22 results were dichotomized (detected vs. not detected) as too few samples contained quantifiable results. Random effects censored regression models were used to test for associations between clinical or microbiome characteristics and cytokine levels (modeled as the response variable). All analyses were conducted in R version 3.6.0.

## RESULTS

### Cohort

Eighty-one subjects consisting of 40 IE and 41 FE provided data and samples during two visits over 2-4 months. Subjects were balanced with respect to age, gender, race, ICS use, and dental care habits (Table 1). Consistent with the VA patient population, most subjects were male. FE phenotype was associated with lower BMI (median 27.9 vs. 30.2, p=0.022), lower FEV1pp (44.0 vs. 52.5, p<0.001), and a higher number of COPD exacerbations in the last 12 months (median 2 vs. 0, mean 2.39 vs. 0). FE were less likely than IE to be current tobacco users (19.5% FE vs. 42.5% IE, p=0.032). Relationships between microbiome measures at each site and clinical factors (exacerbation phenotype, age, FEV1pp, ICS use, pack-years of tobacco use, current tobacco use, toothbrushing frequency, SGRQ score, pathogen detection in sputum samples, or experiencing a COPD exacerbation or use of an antibiotic between study visits) were examined; unless mentioned, these analyses did not reveal an association.

**Table 1.** Subject Baseline Characteristics.

|  | Infrequent<br>Exacerbator<br>(N=40) | Frequent<br>Exacerbator<br>(N=41) | Overall<br>(N=81) | p<br>value * |
| --- | --- | --- | --- | --- |
| Gender, Male (%) | 40 (100) | 40 (97.6) | 80 (98.8) | 1.00 |
| Age, median (IQR) | 69 (5) | 69 (8) | 69 (7) | 0.632 |
| Race, Caucasian white (%) | 37 (92.5) | 40 (97.6) | 77 (95.1) | 0.359 |
| BMI, median (IQR) | 30.2 (7.79) | 27.93 (9.26) | 29.57 (8.46) | 0.022 |
| COPD Severity (%) |  |  |  |  |
| Moderate | 24 (60) | 14 (34.1) | 38 (46.9) |  |
| Severe | 14 (35) | 18 (43.9) | 32 (39.5) |  |
| Very Severe | 2 (5) | 9 (22) | 11 (13.6) |  |
| FEV<sub>1</sub>% predicted, median (IQR) | 52.5 (19) | 44 (17) | 48 (19) | < 0.001 |
| COPD exacerbations in the last 12 months, median (IQR) | 0 (0) | 2 (1) | 1 (2) | < 0.001 |
| COPD hospitalizations in the last 12 months, median (IQR) | 0 (0) | 0 (1) | 0 (0) | < 0.001 |
| Inhaled corticosteroids, Yes (%) | 11 (27.5) | 14 (34.1) | 25 (30.9) | 0.632 |
| Pack-years of smoking, median (IQR) | 41 (23.12) | 50 (23) | 50 (24.5) | 0.059 |
| Current tobacco use, Yes (%) | 17 (42.5) | 8 (19.5) | 25 (30.9) | 0.032 |
| Current alcohol use, Yes (%) | 24 (60) | 32 (78) | 56 (69.1) | 0.096 |
| Brush teeth ≥ once daily (%) <sup>a</sup> | 30 (75) <sup>a</sup> | 30 (75) <sup>a</sup> | 60 (75) <sup>a</sup> | 1.00 |
| SGRQ Score, median (IQR) | 43.62 (10.49) | 52.4 (20.6) | 47.29 (17.84) | 0.074 |

Frequencies and percentages are presented unless specified otherwise. BMI, body mass index; COPD, chronic obstructive pulmonary disease; FEV<sub>1</sub>% predicted, forced expiratory volume in 1 s, percent of predicted value; IQR, interquartile range; SGRQ, St. George's Respiratory Questionnaire
\*A Two-Sample t test was conducted for all continuous variables and a Fisher exact test for all categorical variables
<sup>a</sup>One subject did not provide frequency of brushing teeth

### Alpha Diversity

Alpha diversity was assessed using Shannon diversity, Simpson diversity, and Chao1 metrics. For simplicity, Simpson diversity findings are discussed in detail here (Figure E1), as they are largely consistent with the Shannon and Chao1 diversity findings.

#### Nasal samples

No significant relationships between nasal sample Simpson diversity and any of the evaluated clinical factors were observed.

#### Oral samples

In multiple models of oral wash Simpson diversity, FEV1pp (a model covariate) was consistently associated with increased oral wash Simpson diversity (FE phenotype V1 LR, FEV1pp coefficient estimate [CE] 0.0008, 95% confidence interval [CI, 0.00002, 0.0016], p=0.027, Figure 1A; BV GEE, FEV1pp CE 0.0007, 95% CI [0.0001, 0.0013], p=0.023). In light of this association, we further evaluated potential relationships between FE phenotype, FEV1pp and Simpson diversity. In a model of FE phenotype, FEV1pp, and their interaction, FE phenotype and the interaction of FE phenotype and FEV1pp were associated with oral wash Simpson diversity (interaction BV GEE CE 0.0010, 95% CI [0.00002, 0.0020], p=0.048). Higher SGRQ scores (indicating worse quality of life) were associated with lower oral wash Simpson diversity (V1 LR, CE −0.0007, 95% CI [-0.0013, −0.00011], p=0.018, Figure 1B; and BV GEE, CE −0.0005, 95% CI [-0.0009, −0.0001], p=0.020).

**Figure 1.**
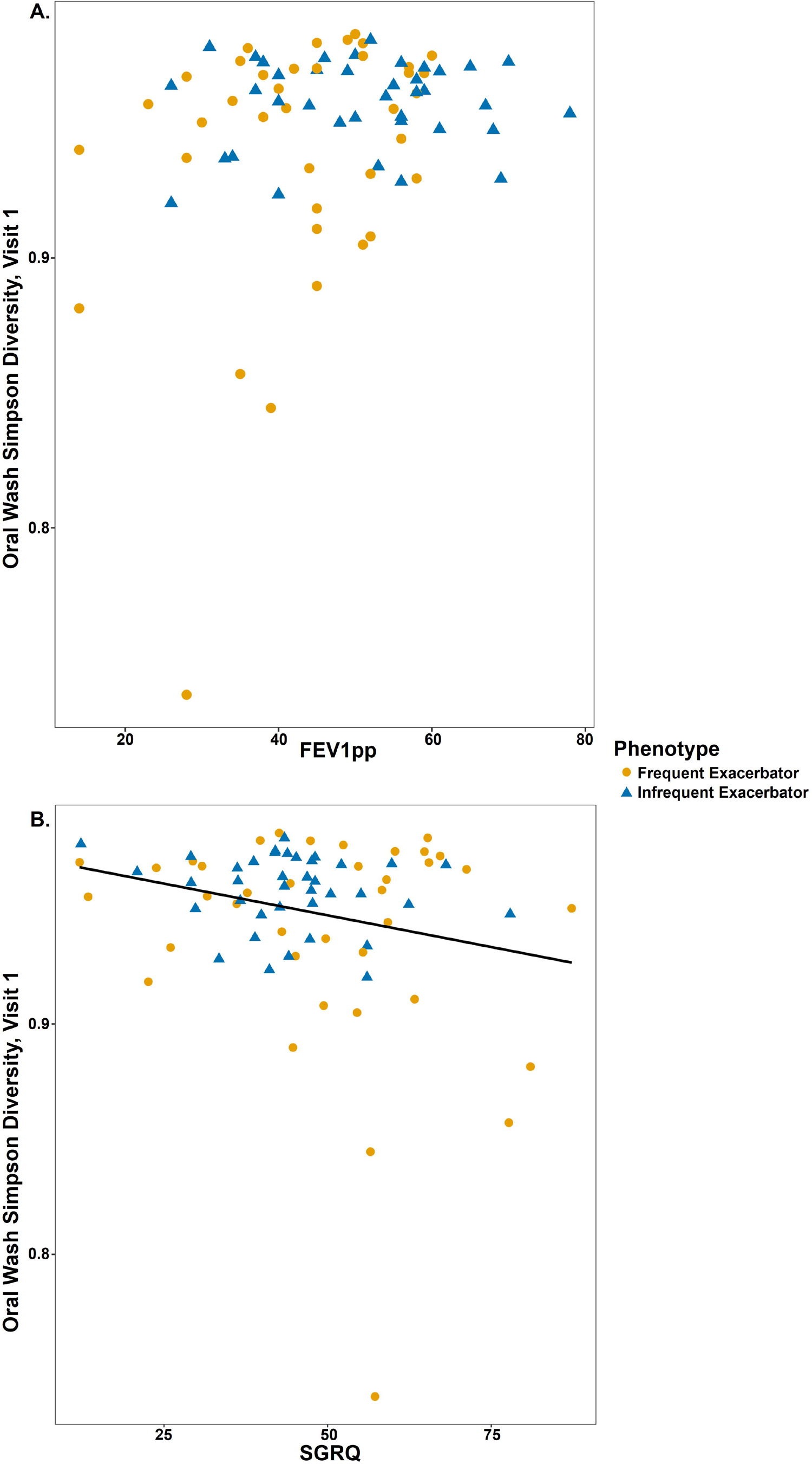
Oral Wash α-diversity is associated with FEV1pp and COPD-related quality of life. A. Oral wash Simpson diversity at visit 1 is associated with FEV1pp (a model covariate) in a model of FE phenotype (FE phenotype V1 LR, FEV1pp CE 0.0008, 95% CI [0.00002, 0.0016], p=0.027. FEV1pp is associated with low Simpson diversity in the adjusted model. B. Oral wash Simpson diversity at visit 1 is associated with SGRQ score at Visit 1 (LR, CE −0.0007, 95% CI [-0.0013, −0.00011], p=0.018). Higher SGRQ scores indicate worse COPD-related quality of life.

#### Sputum samples

The FE phenotype was associated with lower Simpson diversity in sputum at V1 and BV (V1 LR, CE −0.077, 95% CI [-0.15, −0.0048], p=0.041, Figure E2; BV GEE, CE −0.075, 95%CI [-0.12, −0.025], p=0.0031). Older age (another model covariate) was also associated with lower sputum Simpson diversity in a model of exacerbation phenotype (V1 LR, age CE −0.0090, 95% CI [-0.015, −0.0025], p=0.0080, Figures 2a and E3; BV GEE, age CE −0.0074, 95% CI [-0.012. −0.0025], p=0.0030). When exacerbation phenotype, age, and their interaction were included in a model of sputum α-diversity, their interaction was significantly associated with lower Simpson diversity (V1 LR, interaction CE −0.013, 95% CI [-0.024, −0.0024], p=0.020, Figure 2A; BV GEE, interaction CE −0.0099, 95% CI [-0.018, −0.0017], p=0.017). This shows that the association between exacerbation phenotype and sputum Simpson diversity differs based on age.

**Figure 2.**
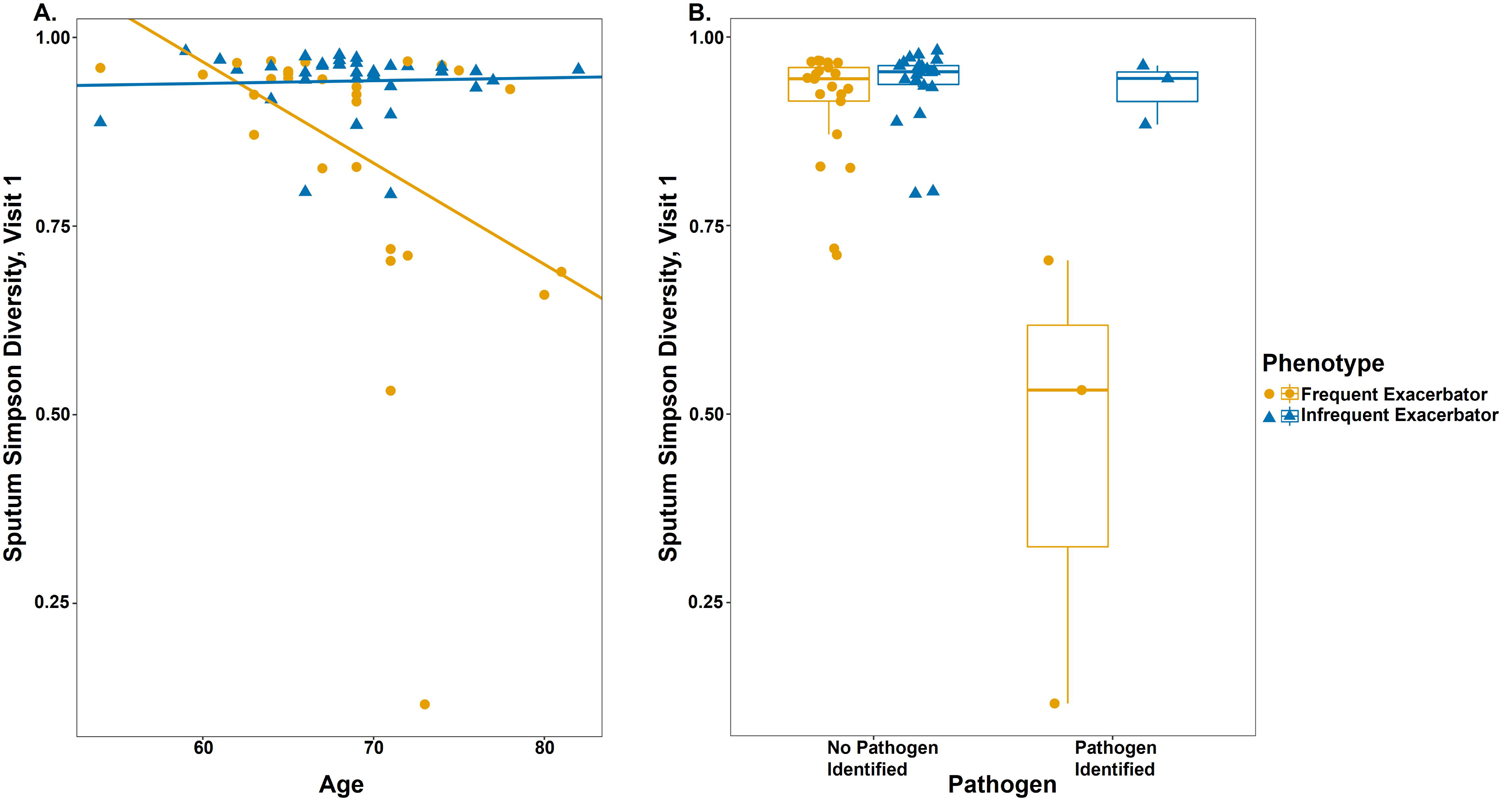
Sputum α-diversity is associated with exacerbation phenotype, age, and culture results. A. Both the FE phenotype and older age were significantly associated with lower Simpson diversity at Visit 1 (LR, CE −0.077, 95% CI [-0.15, −0.0048], p=0.041 and CE −0.0090, 95% CI [-0.015, −0.0025], p=0.0080, respectively). When the interaction of exacerbation phenotype and age was added to the model, only the interaction of age and exacerbation phenotype was significant (LR, CE −0.013, 95% CI [-0.024, −0.0024], p=0.020). The regression lines represent the association between phenotype, age, and Simpson diversity. The significant interaction between age and phenotype indicates that older FE have lower sputum α-diversity than younger FE or older IE. B. Presence of a pathogen in sputum culture was associated with lower sputum Simpson diversity at V1 (LR, CE −0.19, 95% CI [-0.31, −0.074], p=0.0028). When FE phenotype, pathogen, and their interaction were included in the model, only their interaction was significant (V1 LR, CE −0.46, 95% CI [-0.63, −0.30], p<0.001). The association between sputum culture positivity during clinically stable periods and Simpson diversity differs based on exacerbation phenotype.

Sputum samples from 6 subjects (3 FE and 3 IE) were positive for clinically relevant respiratory pathogens (*Moraxella catarrhalis, Haemophilus influenziae*, methicillin-resistant *Staphylococcus aureus*, and *Klebsiella pneumoniae*) at visit 1. Presence of a pathogen was associated with lower sputum Simpson diversity at V1 and BV (V1 LR, CE −0.19, 95% CI [-0.31, −0.074], p=0.0028; BV LMM, CE −0.20, 95% CI [-0.26, −0.14], p<0.001). When FE phenotype, pathogen, and their interaction were included in the model, only their interaction was significant at V1 and BV (V1 LR, CE −0.46, 95% CI [-0.63, −0.30], p<0.001, Figure 2B; BV LMM, CE −0.27, 95% CI [-0.38, −0.15], p<0.001). This shows that the association between sputum culture positivity during clinically stable periods and Simpson diversity differs based on exacerbation phenotype.

In summary, sputum α-diversity is associated with FE phenotype, age, and pathogen detection in sputum culture during clinically stable periods. The association between FE phenotype and decreased sputum α-diversity is also modified by older age or the identification of a pathogen from sputum culture.

#### Alpha diversity over time

Visit 1 values were significantly associated with visit 2 values at all sites (data not shown). Self-reported COPD exacerbation or self-reported use of an antibiotic for any indication in between study visits was not associated with α-diversity at visit 2 (when baseline values were included in the models). Exacerbation phenotype, age, FEV1pp, current tobacco use, pack-years of tobacco use, and SGRQ score were not associated with a change in α-diversity between study visits.

### Beta diversity

Environmental, equipment, and regent control samples were distinct from subject samples (Figure E4). After subsampling to include only subject samples from visit 1, PCoA revealed significant clustering by anatomic site (PERMANOVA, p=0.001 for all pair-wise testing; Figure 3).

**Figure 3.**
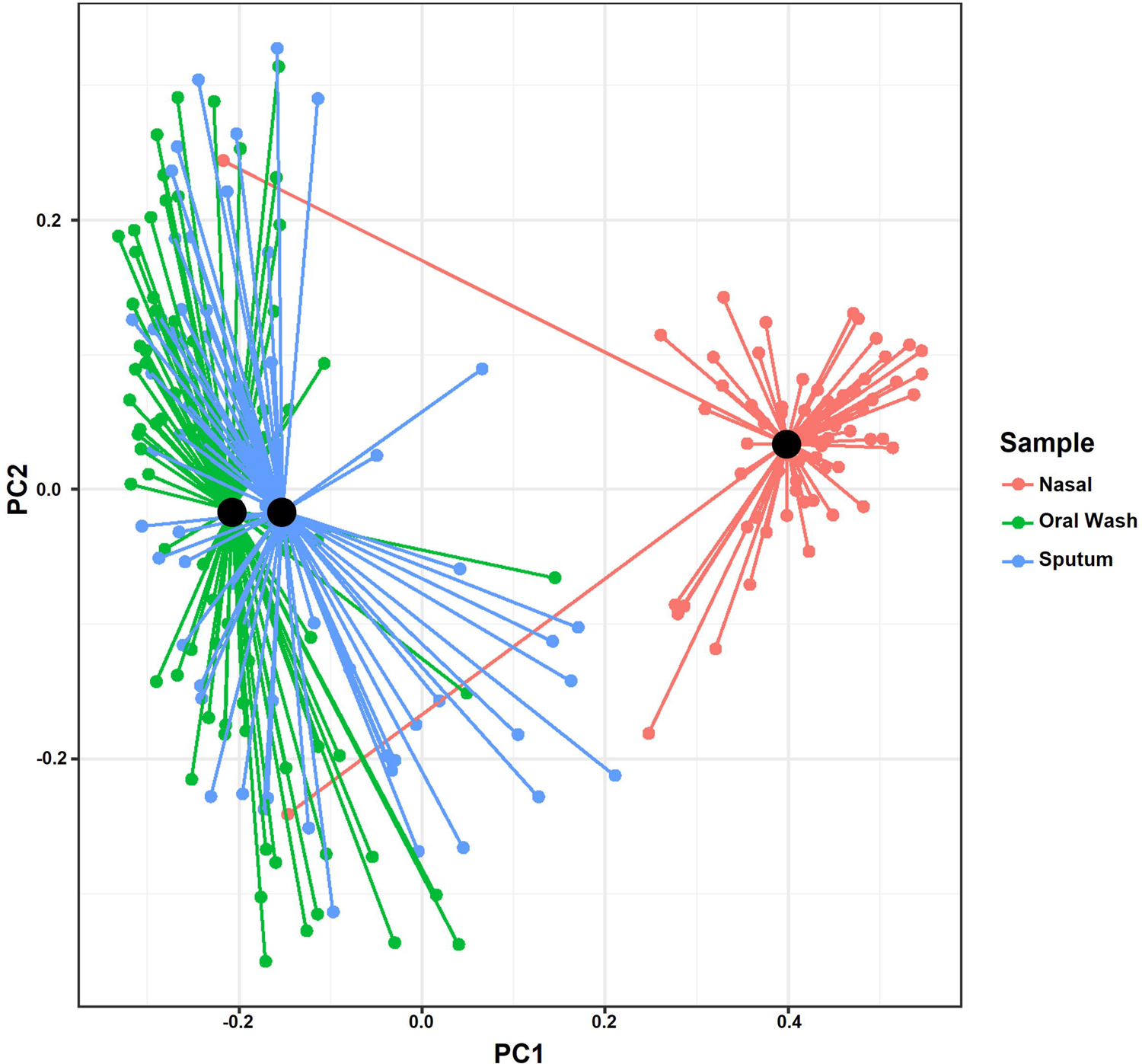
Beta-diversity reveals clustering by anatomic site. Principal coordinate analysis using Bray-Curtis dissimilarity on visit 1 samples demonstrating significant clustering by anatomic site (PERMANOVA, p=0.001 for all pair-wise testing). Centroids are illustrated by black dots, with lines connecting each sample to its corresponding centroid.

#### PERMANOVA analyses

Clustering on PCoA based on exacerbation phenotype and other clinical factors was investigated using PERMANOVA analyses. The analyses were conducted at each anatomic site separately and using visit 1 data, unless noted below. At visit 1, nasal samples clustered based on FEV1pp (PERMANOVA, R^2^=0.029, p=0.033). Several other PERMANOVA results with p<0.10 are provided in Table E1. When visit 2 data were analyzed, sputum samples from participants who reported between-visit antibiotic use for any indication clustered separately from participants who did not report between-visit antibiotic use (PERMANOVA, R^2^=0.029, p=0.049).

#### Beta-diversity over time

Visit 1 samples were compared to corresponding visit 2 samples for all subjects and sites to assess the stability of microbiome composition over time. Among nasal samples, the FE phenotype and antibiotic use between study visits was associated with decreased similarity between paired samples (permutation testing [P], p=0.044 and p=0.032, respectively). There were no associations with oral wash similarity between visits. Among sputum samples, the FE phenotype and experiencing a COPD exacerbation between visits were associated with lower similarity between paired samples (P, p=0.025 and p=0.014, respectively; Figure 4).

**Figure 4.**
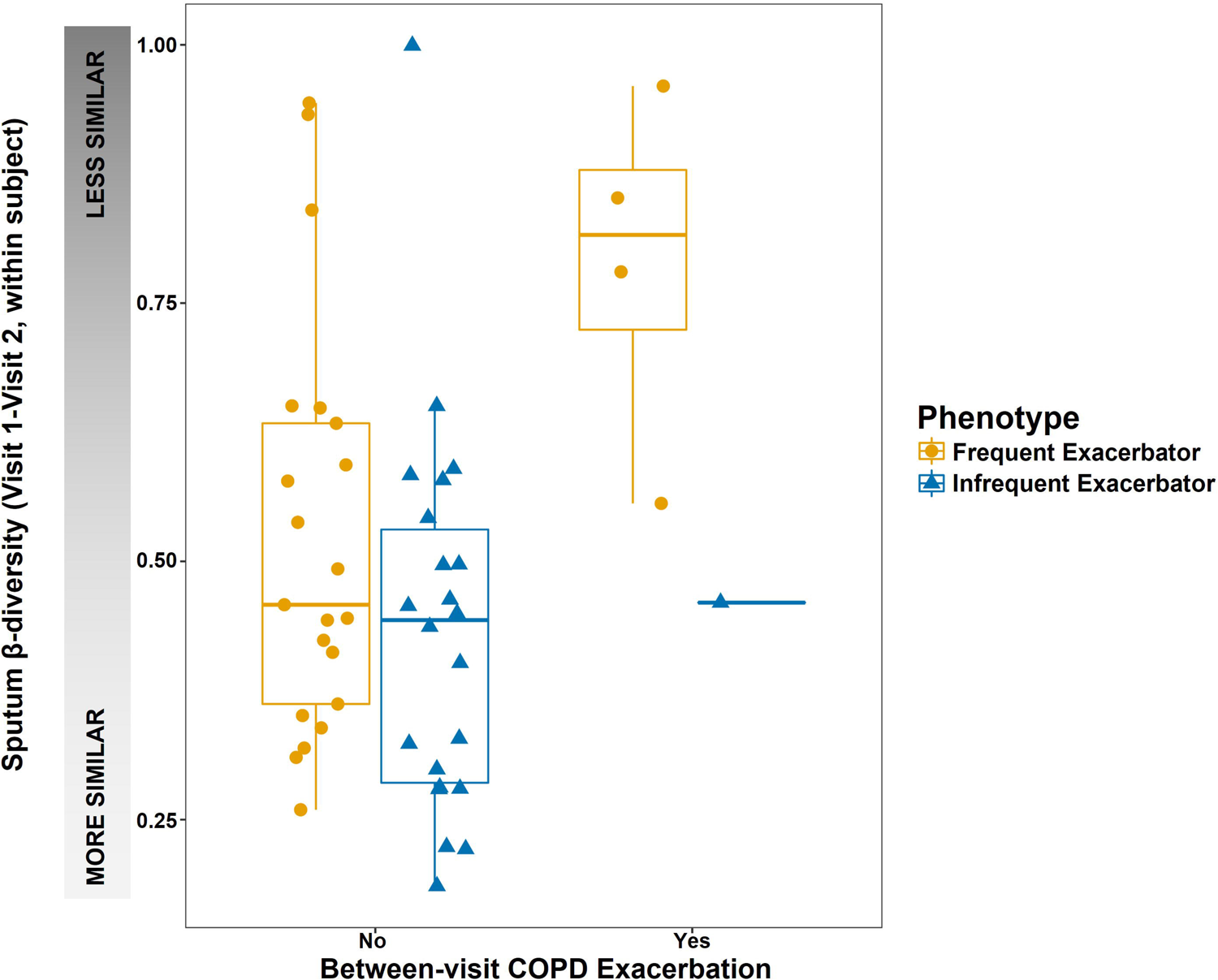
Within-subject microbiome composition stability. Self-report of a COPD exacerbation between study visits corresponded with an increase in microbiome compositional changes (increased β-diversity, or a decrease in sample similarity) compared with subjects who did not experience a COPD exacerbation between study visits.

### Bacterial taxa

We investigated the bacterial taxa present in each sample to determine potential associations with relevant clinical factors (97 tests). Many taxa were associated with clinical site, in accordance with clinical findings and the human microbiome literature. Nasal samples were enriched with *Corynebacterium, Staphylococcus, Cutibacterium,* and *Moraxella* (among others) compared with oral and sputum samples (LR, all p<0.05 following Holm correction; Table E2). Oral and sputum samples were enriched with *Veillonella, Rothia, Prevotella, Streptococcus*, and *Haemophilus* (among others) when compared with nasal samples (LR, all p<0.05 following Holm correction; Table E3). Across all anatomic sites, *Mannheimia* abundance was positively associated with age (LR, p=0.0014); *Mogibacterium* abundance was lower among FE compared to IE (p=0.029); *Leuconostoc* abundance was negatively associated with FEV1pp (p=0.020); *Bulleidia* abundance was higher among current tobacco users (p=0.013); and *Pseudomonas* abundance was positively associated with pack-years of tobacco use (p<0.0001).

### Sputum cytokine analyses

Sputum sample cytokine levels were tested for an association with clinical factors and sputum microbiome characteristics. Cytokines were chosen for analysis based on prior reported associations with culture or microbiome results, and analyses of the 7 evaluable cytokines are provided here. Samples from all available visits were analyzed using GEE or random effects censored regression models, as appropriate, accounting for visit.

#### Cytokines associated with clinical characteristics

None of the clinical factors (exacerbation phenotype, age, FEV1pp, pack-years of tobacco exposure, current tobacco use, or SGRQ score) were associated with cytokine levels in a random effects censored regression model.

#### Cytokines associated with Alpha diversity (Simpson)

Three sputum cytokines were associated with sputum sample α-diversity on univariate analysis. IL-17A levels were negatively associated with Simpson diversity, while IL-6 and IL-8 were positively associated with Simpson diversity (random effects censored regression model, IL-17A CE −1.3, 95% CI [-2.2, −0.50], p=0.012; IL-6 CE 6.0, 95% CI [2.2, 9.8], p=0.012; and IL-8 CE 4.2, 95% CI [1.0, 7.4], p=0.0496; respectively; Figure 5). IL-22, IL-5, IL-1β, and LTB4 were not associated with Simpson diversity.

**Figure 5.**
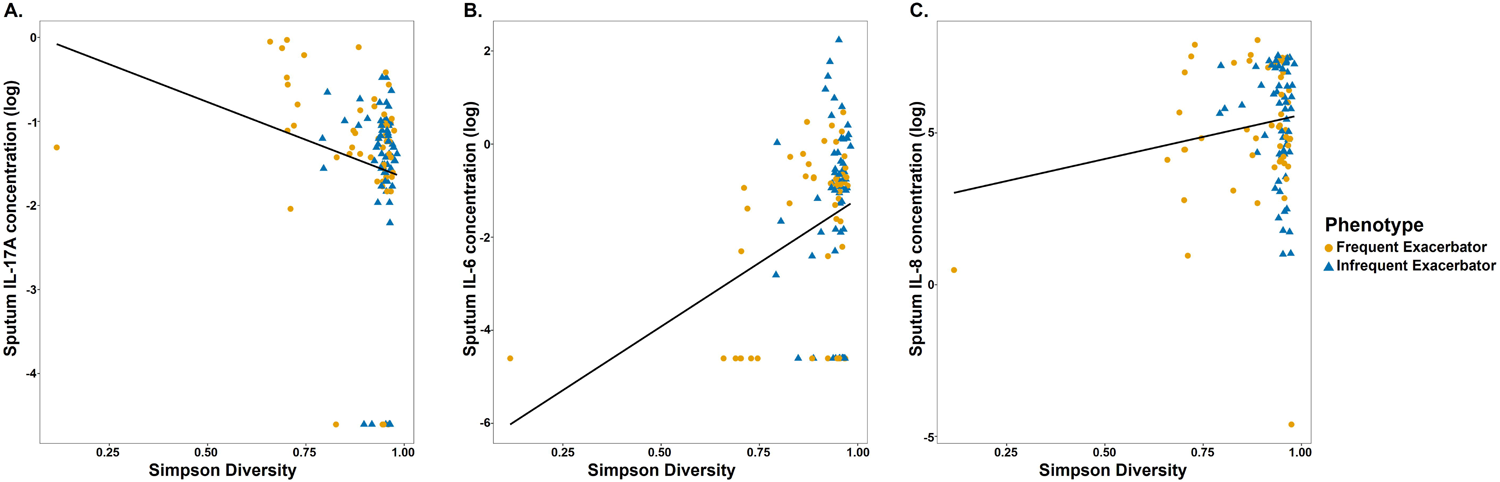
Multiple sputum cytokines are associated with α-diversity. Increased sputum concentrations of IL-17A were associated with decreased Simpson diversity in the sputum, while increased sputum concentrations of IL-6 and IL-8 were associated with increased Simpson diversity (random effects censored regression model, IL-17A CE −1.3, 95% CI [-2.2, −0.50], p=0.012; IL-6 CE 6.0, 95% CI [2.2, 9.8], p=0.012; and IL-8 CE 4.2, 95% CI [1.0, 7.4], p=0.0496; respectively).

#### Cytokines associated with positive bacterial sputum culture

Presence of typical COPD pathogens in sputum samples, such as *Streptococcus*, *Moraxella,* or *Haemophilus*, have been associated with exacerbation phenotype, increased inflammation, and decreased α-diversity. We found that IL-22, IL-17A, and IL-5 levels were positively associated with *Moraxella* abundance (GEE with Holm correction, IL-22 CE 10.26, 95% CI [3.96, 16.55], p=0.027; random effects censored regression model with Holm correction, IL-17A CE 2.01, 95% CI [1.02, 2.99], p=0.0014; and IL-5 CE 1.87, 95% CI [0. 93, 2.81], p=0.002, respectively; Figure 6). There were no significant associations with other cytokines or the genera *Streptococcus* or *Haemophilus*.

**Figure 6.**
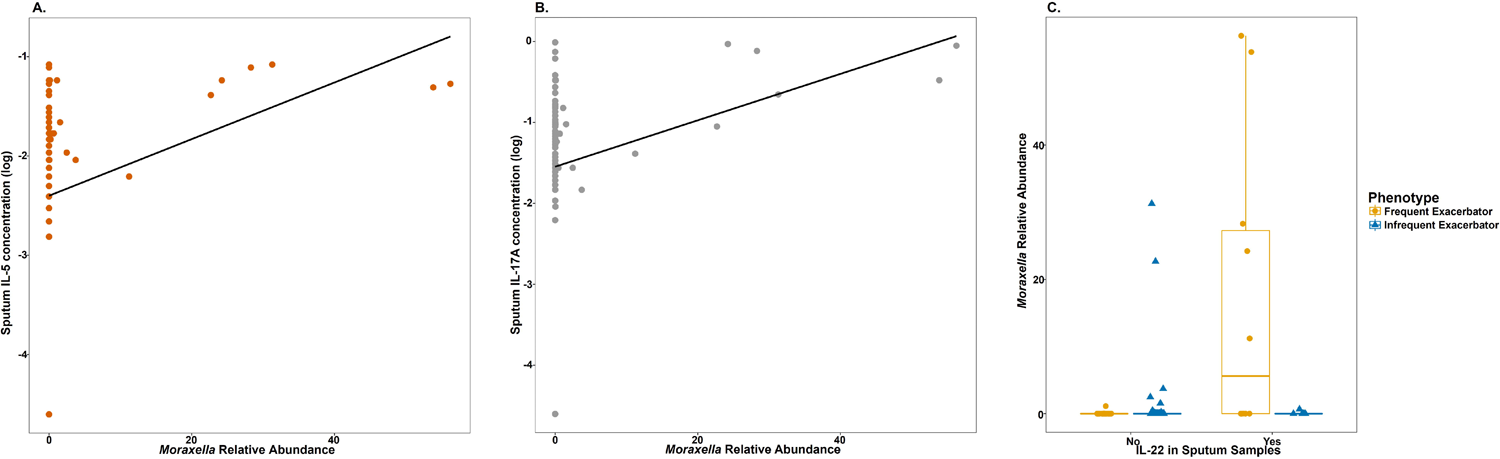
Sputum cytokines are associated with *Moraxella* abundance. Increased sputum concentrations of IL-22, IL-17A, and IL-5 were correlated with increased abundance of *Moraxella* in sputum samples (GEE with Holm correction, IL-22 CE 10.26, 95% CI [3.96, 16.55], p=0.027; random effects censored regression model with Holm correction, IL-17A CE 2.01, 95% CI [1.02, 2.99], p=0.0014; and IL-5 CE 1.87, 95% CI [0.93, 2.81], p=0.002, respectively).

## DISCUSSION

Our case-control longitudinal observational study of the upper airway and sputum microbiome during periods of clinical stability identified low sputum microbiome α-diversity as a key feature of the COPD frequent exacerbator phenotype. In turn, low sputum microbiome α-diversity was associated with airway bacterial colonization and lung inflammation, two other characteristics consistent with the increased morbidity and mortality associated with the frequent exacerbator phenotype. Additionally, the frequent exacerbator phenotype was associated with decreased microbiome compositional stability (increased β-diversity) on longitudinal sputum sampling. These findings suggest that low α-diversity and an unstable sputum microbiome are key features of the frequent exacerbator phenotype.

In addition to our findings on the frequent exacerbator phenotype, we also detected associations between the sputum, oral, or nasal microbiome and age and COPD-related symptom severity. Also, among the small number of subjects who experienced an exacerbation between study visits, we observed compositional changes (increased β-diversity) in the microbiome following the occurrence of a COPD exacerbation. Our study determined that the frequent exacerbator phenotype is associated with low sputum microbiome α-diversity. Low α-diversity among FE has been found by many— but not all—investigators, possibly related to differences in how exacerbation frequency was analyzed [20, 22–26, 28, 29]. In the case of the sputum microbiome, this homogeneity was enhanced among older participants and among participants with airway bacterial colonization. This phenotype also correlated with increased *Mogibacterium*, an oral taxon associated with oral inflammation and stable COPD [30, 31].

In this study and others, older age was associated with decreased α-diversity among sputum samples [23, 28]. Here, we also determined that older age is associated with a further decrease in α-diversity among FE. Although aging is associated with an increased likelihood of COPD diagnosis—as well as increased susceptibility to lung infections and declines in lung function—the associations between aging and the microbiome are relatively understudied. The aging COPD lung microbiome and the gut-lung axis may exist at a juncture between declining lung function, immunosenescence, nutritional changes, and increased antibiotic use [32]. Our work suggests that age itself may influence the COPD lung microbiome, possibly via mechanisms independent of typical factors (such as COPD exacerbations) that are known to influence the lung microbiome.

Our analysis of associations between the microbiome and tobacco use encompassed both pack-years of tobacco use as well as current (vs. former) tobacco use status. Consistent with many of the previous studies [33–35], which identified more tobacco-associated microbiome changes of the upper vs. lower airway, we identified that greater pack-years of tobacco exposure were associated with increased *Pseudomonas* abundance, while current (vs. former) tobacco use was associated with increased *Bulleidia* abundance, primarily in the oropharynx. *Bulleidia* has previously been associated with tobacco use and lung cancer [36, 37].

The present study also evaluated the longitudinal stability of the upper and lower airway microbiome. Although microbiome findings were generally similar on repeated sampling (low β-diversity), we identified several scenarios in which β-diversity was increased. The sputum microbiome of FE exhibited decreased similarity (increased β-diversity) compared with IE. Subjects who experienced a COPD exacerbation between study visits also exhibited decreased compositional similarity vs. those who did not report interim exacerbations.

We also found significant differences in sputum cytokine levels which associated with sputum microbiome α-diversity, but not related to exacerbation phenotype, COPD severity, age, tobacco use, or COPD-related quality of life. Low sputum α-diversity was associated with increased concentrations of sputum IL-17A and decreased concentrations of IL-6 and IL-8. *Moraxella* abundance in the sputum microbiome was also associated with increased concentrations of IL-22, IL-17A, and IL-5. IL-17A, IL-22, and IL-6 are key mediators of a Th17 response, often at mucosal sites [38]. IL-8 is a neutrophil chemoattractant. IL-5 is involved in Th2 responses and eosinophil recruitment. In prior COPD studies, elevated sputum IL-6 and IL-8 levels have been associated with tobacco use, lower FEV1pp, frequent exacerbations, and acute exacerbation (vs. clinical stability) [8, 39–42]. Our finding of a positive association between IL-6 and IL-8 levels and α-diversity is somewhat unexpected, but it is possible that the acute rise in IL-6 and IL-8 observed during exacerbations is not reflected in our samples, which were collected during exacerbation-free intervals. Furthermore, few investigators have assessed the sputum microbiome—specifically low α-diversity—in relationship to inflammatory cytokines.

Our manuscript has several strengths. We used well-defined FE and IE phenotypes consistent with GOLD guidelines [27], allowing us address associations between the microbiome and exacerbation phenotype. We deferred all study visits for 1 month following a COPD exacerbation or antibiotic use for any reason, in order to focus on the microbiome during periods of clinical stability. This approach minimizes, to the extent possible in an observational study of COPD, the influence of recent antibiotic or systemic steroid use on our microbiome findings. Lastly, our longitudinal approach allowed us to assess the stability of microbiome composition in relationship to exacerbation phenotype and recent COPD exacerbations. Our use of sputum inflammatory cytokines, in the context of the microbiome findings, identified clinical correlates of our microbiome findings.

Despite these strengths, our study had several relative weaknesses. We are unable to assess the influence of sex on the microbiome, as our single-center study was conducted at a Veterans Affairs hospital with a limited female population. Sputum samples may be contaminated by saliva during expectoration and therefore may not reflect only the lower airway microbiome. Despite this potential limitation, we note that most of our key microbiome findings were identified only in the sputum microbiome and not identified in the oral microbiome. This suggests that sputum analysis can identify microbiome associations unique to the lower airways, despite potential upper airway contamination.

In conclusion, we found that FEs exhibit lower sputum microbiome α-diversity, which is enhanced by older age or bacterial colonization of the airways. Sputum microbiome α-diversity is a significant correlate of lung inflammation. The sputum microbiome composition of FEs changes more over time when compared to the compositional stability of IEs.

## Data Availability

All data produced are available online at NCBI.

## ACKNOWLEDGEMENTS

The authors wish to thank Ms. Susan Johnson, LPN, for assistance with study recruitment. This material is based upon work supported by the Department of Veterans Affairs, Veterans Health Administration, Office of Research and Development (Clinical Sciences Research and Development). The views expressed in this article are those of the authors and do not necessarily reflect the position or policy of the Department of Veterans Affairs or the United States government. This publication was supported by Grant Number 1UL1RR033183 from the National Center for Research Resources (NCRR) and by Grant Number 8 UL1 TR000114-02 from the National Center for Advancing Translational Sciences (NCATS) of the National Institutes of Health (NIH) to the University of Minnesota Clinical and Translational Science Institute (CTSI). Its contents are solely the responsibility of the authors and do not necessarily represent the official views of the CTSI or the NIH. The University of Minnesota CTSI is part of a national Clinical and Translational Science Award (CTSA) consortium created to accelerate laboratory discoveries into treatments for patients.

## Take home message

Low sputum microbiome α-diversity and decreased compositional stability (increased β-diversity) are associated with the COPD frequent exacerbator phenotype. Low sputum microbiome α-diversity is also associated with lung inflammation.

## Authors’ contributions

Conception and Design: A.A.P., C.S.R., and C.H.W. Data Acquisition: A.A.P., S.W.H., and A.Z. Data analysis: A.A.P., T.W., C.S.R., and C.H.W. Data interpretation: A.A.P., T.W., C.S.R., and C.H.W. Drafting the manuscript: A.A.P. Revision and final approval of the version to be published: All authors.

---

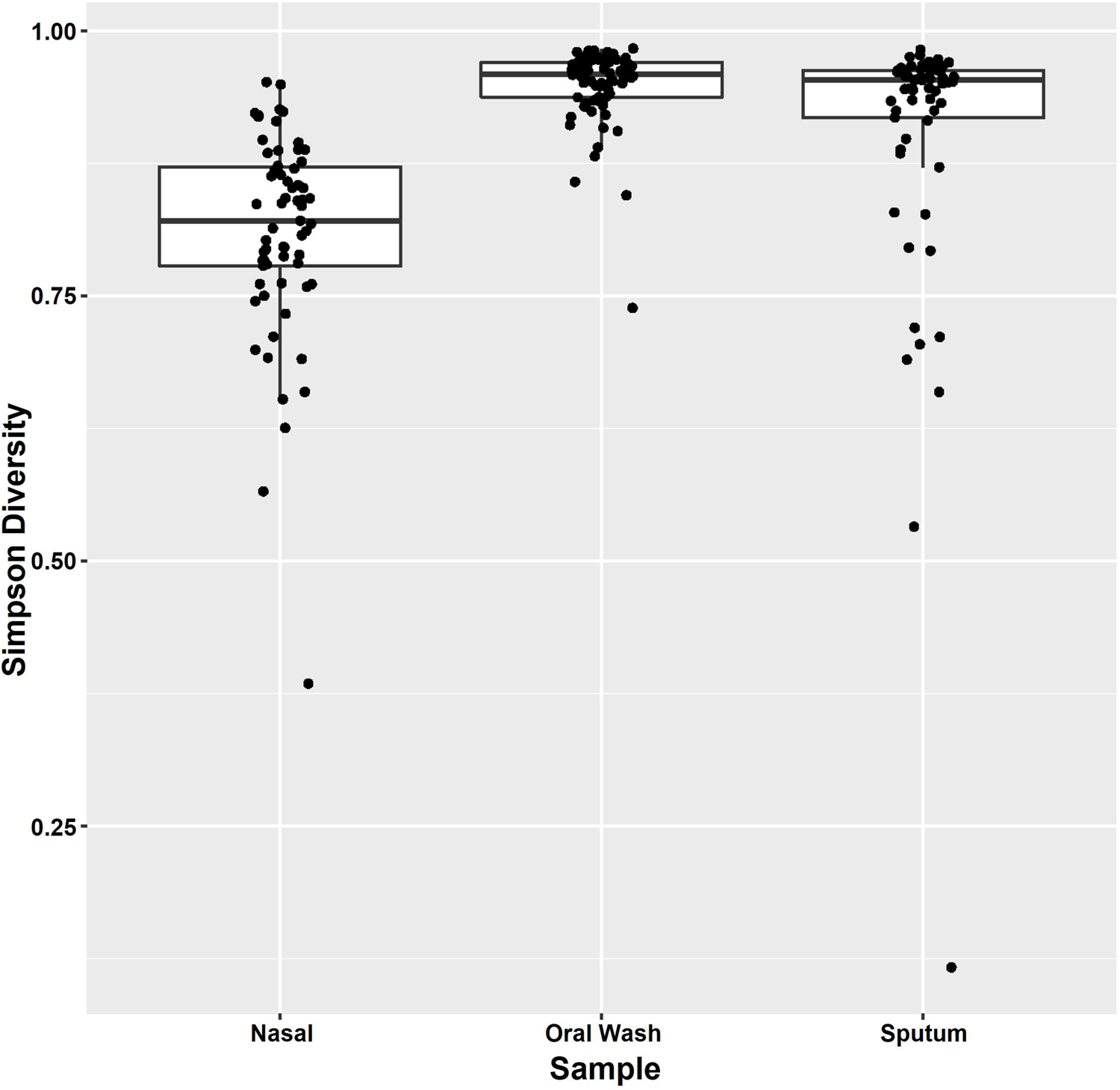

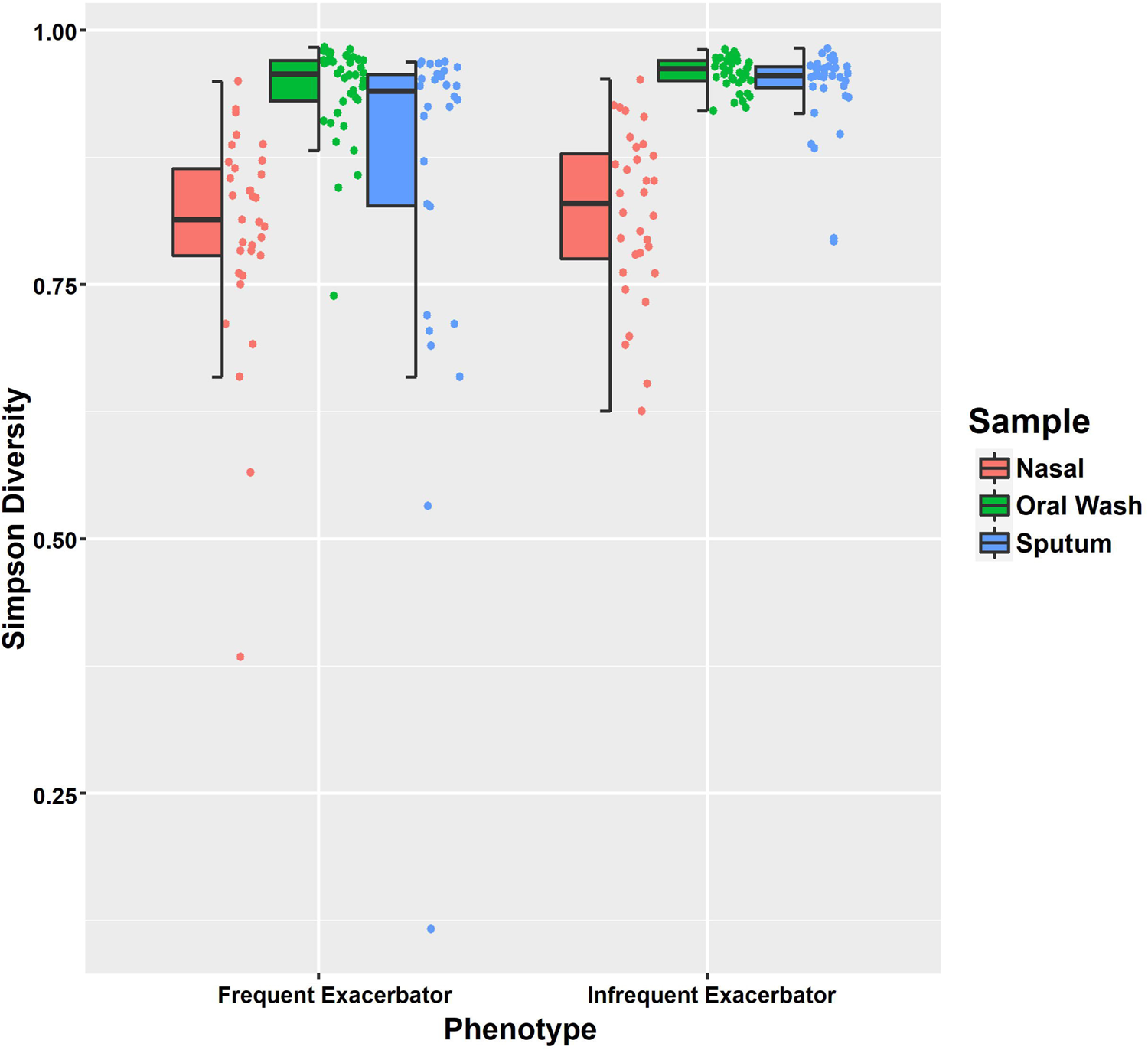

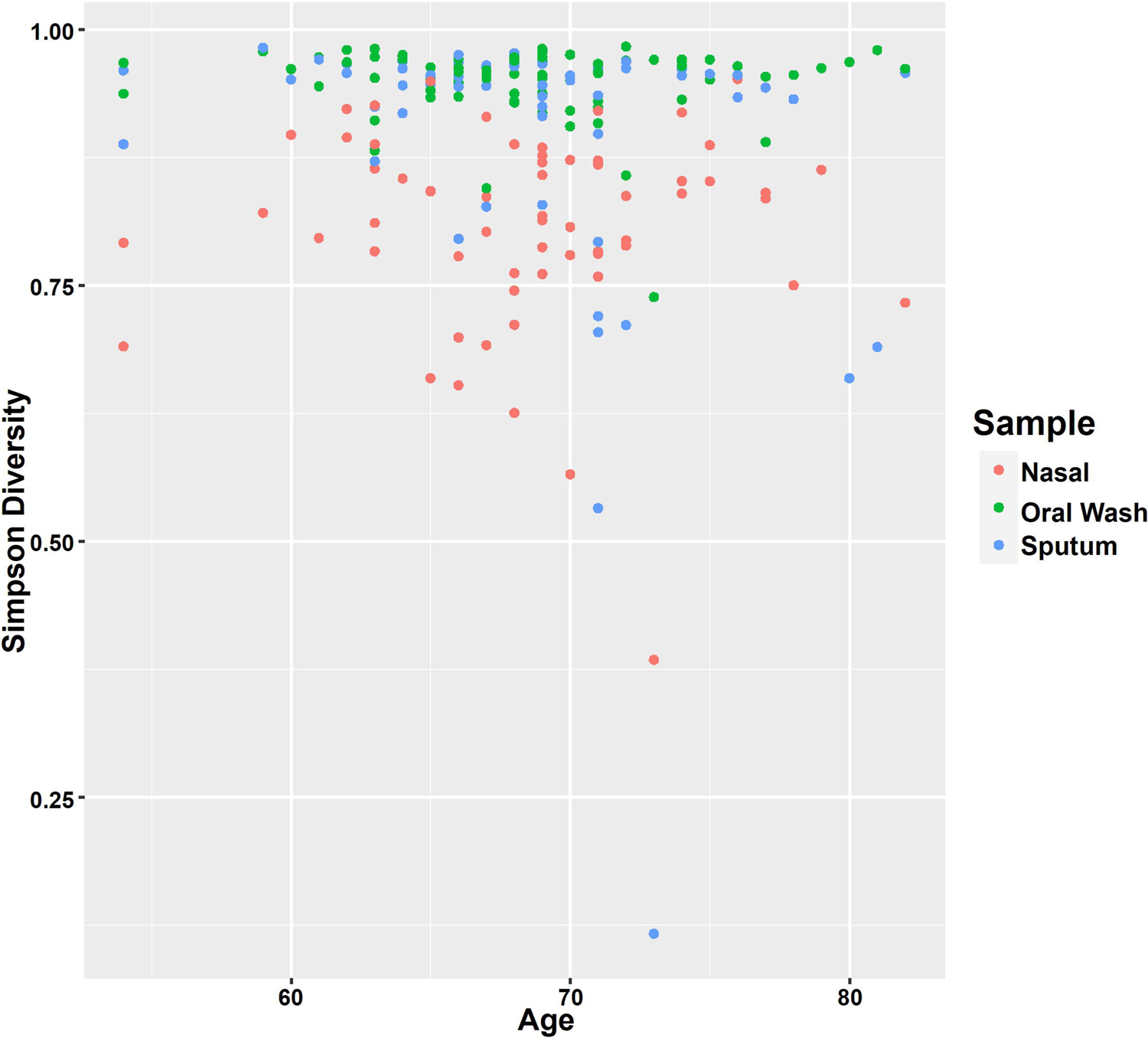

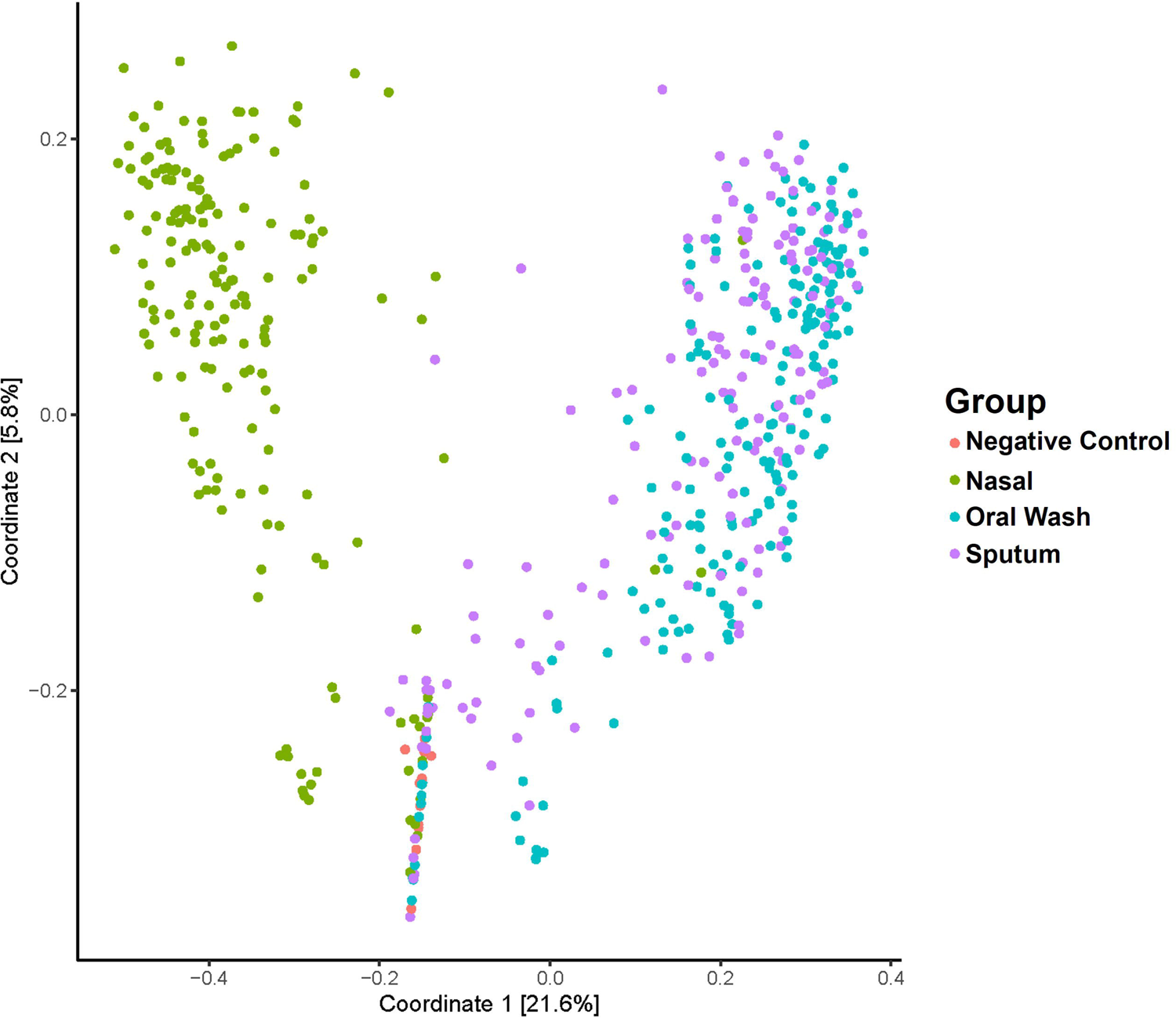

