## Supplementary Information for "Sputum microbiome α-diversity is a key feature of the COPD frequent exacerbator phenotype"

**Sputum microbiome α-diversity is a key correlate of the COPD frequent exacerbator phenotype and lung inflammation**

**ONLINE DATA SUPPLEMENT**

**Supplementary Methods:**

**Study design and recruitment.** We conducted a case-control longitudinal observational study of exacerbation phenotype and characteristics of the upper airway and sputum microbiome. All participants were recruited from the Minneapolis VA Medical Center (MVAMC) and must have been age 40 or older, have FEV1/FVC <70% and post-bronchodilator FEV1<70% predicted, have at least a 10 pack-year history of tobacco use, have no history of asthma or lung lobectomy, no use of an antibiotic for any indication in the past 1 month, and have a room air oxygen saturation of >89%. Frequent exacerbators (FE) must have had ≥ 1 exacerbation requiring hospital admission or emergency department visit in the last 12 months (defined as the use of an antibiotic or corticosteroid for a respiratory indication), while infrequent exacerbators (IE) must not have had any exacerbations in the prior 24 months. Our definition of FE is consistent with the most recent GOLD guidelines.[1] Among the 41 FE, at enrollment 9 reported 1 exacerbation, 17 reported 2 exacerbations, and 15 reported 3 or more exacerbations in the last 12 months. To focus on the microbiome during clinical stability, recruitment and sampling were deferred until the subject had recovered for at least 1 month from the most recent exacerbation. The protocol was approved by the Minneapolis VA IRB (#4541-B).

**Study procedures.** After signing the informed consent form during visit 1, participants provided their medical history, underwent spirometry, and completed the St. George’s Respiratory Questionnaire (SGRQ). Oral wash samples were obtained by swishing 10 mL of sterile water in their mouths for 30 seconds. Nasal samples were obtained by nasal swab (nylon flocked swab, COPAN Diagnostics, Murrieta, CA, USA; 15 seconds each nostril). Sputum was induced with hypertonic saline and divided for further studies. Oral wash and sputum samples were weighed. 1 mL of the sputum sample was sent to the clinical microbiology lab for routine sputum gram stain and culture. The remainder of the sputum sample, as well as the oral and nasal samples, were frozen at -70 ºC prior to further processing. All participants returned for a second study visit approximately 2 months following visit 1, where they provided information on any interim COPD exacerbations, repeated the SGRQ, and provided oral, nasal, and sputum samples as at visit 1. Visit 2 was deferred (for up to 4 months following visit 1) if the participant reported any COPD exacerbations in the 1 month prior to the visit. Environmental and reagent contamination controls consisted of unused nasal swabs and unused sterile saline which were processed alongside samples as below.

**Sample processing.** All samples were extracted using the MO BIO PowerSoil DNA Isolation Kit (QIAGEN, Germantown, MD) with modifications as noted below. Nasal swabs were thawed and transferred to a PowerBead tube followed by addition of PowerSoil solution C1 and incubation at 70 °C for 15 minutes. Oral wash samples were centrifuged at 13,000g for 15 minutes to pellet the sample, then resuspended in PowerBead tube buffer solution and transferred back to the tube. Sputum samples were incubated at 37 °C with an equal volume of Sputolysin (Millipore-Sigma, Darmstadt, Germany) for 15 minutes, then divided equally for cytokine analyses and sequencing. The portion used for sequencing was pelleted at 13,000g for 15 minutes and resuspended in PowerBead tube solution for transfer into the tube. Once placed in the PowerBead tubes, the samples were extracted following the manufacturer’s instructions.

**16S rRNA gene quantification and sequencing.** Extracted DNA from each sample was submitted to the University of Minnesota Genomics Center for 16S rRNA gene quantification and 16S rRNA gene V4 MiSeq sequencing. 16S rRNA gene quantification was accomplished with droplet digital PCR (ddPCR) on the Bio-Rad QX200 instrument using a standard curve created with serial dilutions of *E. coli* template DNA tested in triplicate. PCR was accomplished using the 2X EvaGreen SuperMix (Bio-Rad) and 16S rRNA V4 primers (GTGCCAGCMGCCGCGGTAA, GGACTACHVGGGTWTCTAAT). ddPCR reactions were performed as follows (ramp rate of 2 °C/second): 95 °C for 5 minutes; followed by 45 cycles of 95 °C for 30 seconds/60 °C for 90 seconds; then 4 °C for 5 minutes followed by 90 °C for 5 minutes. Libraries were prepared for 16S rRNA V4 marker gene sequencing on the MiSeq instrument using dual-indexing as described previously.[2]

**16S rRNA sequence processing**. Sequences were trimmed for sequencing artifacts and primers with cutadapt (https://cutadapt.readthedocs.io/en/stable/#). Further quality trimming, error correction, chimeric read removal, and ASV table creation were done with DADA2.[3] Taxonomic identification was done with DADA2 using RDP Taxonomy 18.[4] ASV table processing was done using R as described here: https://github.com/trevorjgould/MarkerGeneAnalysis. α-diversity metrics were obtained using OTUtable (chao1) and Vegan (Shannon diversity index, Simpson diversity index). β-diversity was assessed using the Bray-Curtis matrix with CLR transformation followed by principal component analysis as outlined here: https://github.com/trevorjgould/MarkerGeneAnalysis/blob/master/R/diversity.R. Initial analyses were performed with negative control samples included in order to understand reagent and equipment contaminants, as shown in Figure E4. Based on these analyses, no samples or sequences were removed from the dataset. Subsequent analyses, as reported in the main manuscript, were conducted after excluding all control samples from the dataset.

**Sputum culture results.** The clinical microbiology laboratory at MVAMC performed routine sputum Gram stain and aerobic culture on sputum samples. Samples with >10 squamous cells per high-powered field and <25 polymorphonuclear cells per high-powered field on Gram stain did not undergo culture. Samples that did not meet these criteria underwent aerobic culture and results were reported per clinical protocols. Any sample reported as growing a named bacterial organism was categorized as growing a clinically-relevant organism.

**Cytokine analyses.** Sputum samples (0.5 mL) were submitted to the University of Minnesota Cytokine Reference Laboratory for determination of LTB4 and IL-22 (via ELISA), IL-8, IL-6, IL-1β, IL-5, IL-17A, GM-CSF and TNF-α (via Luminex, all R&D Systems, Minneapolis, MN USA).

**Statistical analyses.** All analyses were conducted in R version 3.6.0. Counts and percentages were used to summarize categorical variables, while medians and interquartile ranges were used to summarize continuous variables. Univariate tests of association of continuous variables with a binary variable were conducted using 2 sample t-tests. Fisher’s exact test was used to conduct univariate tests between categorical variables. Data presented is from visit 1 only (V1) or both visits (BV) as outlined in the text. Linear regression (LR), including random effects censored regression models, linear mixed models (LMM), and generalized estimating equations (GEE), were used to test for associations between variables. Analyses of α-diversity metrics were adjusted for age, FEV1pp, BMI, current tobacco use, and current alcohol use. These clinical factors were chosen based on their correlation with phenotype (Table 1). To increase power, data from the 2 visits was pooled when relevant. β-diversity was assessed via the Bray-Curtis dissimilarity metric and illustrated via principal coordinates analysis (PCoA). PERMANOVA analyses of β-diversity were performed for each anatomical site individually to ensure all datapoints were independent. To compare the within-subject similarity between visits across categorical clinical variables (exacerbation phenotype, current tobacco use, between-visit antibiotic use, and between-visit COPD exacerbation), permutation tests were performed with 1,000 iterations. The p-values obtained were used to determine if the categorical clinical variable was associated with changes in within-subject similarity between visit 1 and visit 2. A p-value <0.05 was deemed statistically significant, unless otherwise noted and all tests are 2-sided. For the taxonomic analysis, genera found in <10% of samples were removed from the dataset prior to analysis. Tests of association between bacterial taxa and clinical characteristics used Holm’s procedure to control the family-wise error rate at 5% for each clinical characteristic. Random effects censored regression models were used to test for associations between clinical or microbiome characteristics and cytokine levels. GM-CSF and TNF-α were removed from the analysis because there were insufficient quantifiable results to conduct a continuous analysis.

**Supplementary Results:.**

*α-diversity analyses.* Simpson diversity index, Shannon diversity index, and Chao1 diversity index were reported for each sample and correlations with clinical factors were assessed as described above. Nasal samples were less diverse than sputum samples, which were less diverse than oral wash samples for all α-diversity metrics (Figure E1, p<0.05 for all pair-wise comparisons of Simpson diversity with Bonferroni adjustment).

*β-diversity analyses*. Bray-Curtis dissimilarity and principal coordinate analysis was used to illustrate microbial composition and between-sample similarity. Samples were split by sampling site and PERMANOVA analyses using Bray-Curtis dissimilarity were used to identify clustering based on clinical factors. To compare within-subject visit 1-visit 2 dissimilarity, permutation tests (1,000 iterations) were performed to determine if clinical factors were associated with changes in within-subject similarity.

*Taxonomic composition*. Among visit 1 samples, only genera that were present in at least 10% of samples were used for this analysis (97 genera). Linear regression models with log(reads+1) as the response variable and relevant clinical factors as the explanatory variables were used to detect associations. The Holm method was used to adjust p-values for multiple hypothesis testing.

*Cytokine analysis*. Sputum samples from visit 1 and visit 2 were analyzed. Data on GM-CSF was not analyzed because it was detected below the lower limit of quantitation in all samples. TNF-α was not analyzed because it was undetected in 5 samples and detected below the lower limit of quantitation in the other samples. IL-22 was detected above the lower limit of quantitation in 28 samples, but undetected in the remainder of samples. Therefore, IL-22 data were dichotomized into “detected” or “not detected” before GEE analysis. For IL-17A, IL-1β, IL-5, IL-6, IL-8 and LTB4, samples with cytokine levels below the lower limit of quantitation were censored and values were log transformed before using random effects censored regression model. All models also accounted for the plate used.

**Supplementary Results:**

We evaluated our results to determine whether there were associations between extraction kit used and our findings. Our analyses of 16S copy number data and α-diversity indices showed no relationship between kit used for either of these results (data not shown).

**Supplementary Figures.**

**Figure E1. Simpson diversity by site.** Nasal samples were less diverse than sputum samples, which were less diverse than oral wash samples (p<0.05 for all pair-wise comparisons with Bonferroni adjustment).

**Figure E2. Simpson diversity by site and phenotype.** The FE phenotype was associated with decreased Simpson diversity in sputum at V1 (V1 LR, CE -0.077, 95% CI [-0.15, -0.0048], p=0.041). Exacerbation phenotype was not associated with nasal or oral sample Simpson diversity.

**Figure E3. Simpson diversity by age and site**. Nasal, oral wash, and sputum sample Simpson diversity at visit 1 was plotted by subject age. Older age was associated with decreased Simpson diversity in sputum at V1 (V1 LR, CE -0.0090, 95% CI [-0.016, -0.0025], p=0.0080). Analysis of the nasal and oral wash sites was not significant.

**Figure E4. PCoA plot including negative control samples.** Negative control samples (environment, reagent, and equipment controls) clustered at bottom, while nasal samples clustered at left and oral wash and sputum samples clustered at right.

**Supplementary Tables.**

**Table E1. PERMANOVA Analyses.**

| **Characteristic** | **Nasal** | | **Oral Wash** | | **Sputum** | |
| --- | --- | --- | --- | --- | --- | --- |
|  | **R^2^** | **p-value** | **R^2^** | **p-value** | **R^2^** | **p-value** |
| Phenotype |  |  | 0.019 | 0.082 | 0.023 | 0.080 |
| FEV1pp | **0.029** | **0.033** |  |  | 0.024 | 0.057 |
| Pack-years of tobacco use |  |  | 0.019 | 0.070 |  |  |
| SGRQ score | 0.024 | 0.077 |  |  |  |  |
| Interim COPD exacerbation |  |  | 0.023 | 0.075 | 0.029 | 0.051 |
| Interim antibiotic use (any indication) |  |  |  |  | **0.029** | **0.049** |

**Table E2. Taxonomic analyses by anatomic site.**

| **Corresponding Genus** | **Adjusted p value for overall effect of Site^a^** | **Coefficient for Oral Wash** | **Adjusted p value for Oral Wash^b^** | **Coefficient for Sputum** | **Adjusted p value for Sputum^c^** |
| --- | --- | --- | --- | --- | --- |
| *Cutibacterium* | < 0.0001 | -4.8497 | < 0.0001 | -5.1041 | < 0.0001 |
| *Staphylococcus* | < 0.0001 | -5.3267 | < 0.0001 | -5.7569 | < 0.0001 |
| *Anaerococcus* | < 0.0001 | -4.5694 | < 0.0001 | -5.0833 | < 0.0001 |
| *Finegoldia* | < 0.0001 | -3.8883 | < 0.0001 | -3.8422 | < 0.0001 |
| *Veillonella* | < 0.0001 | 4.8701 | < 0.0001 | 4.7702 | < 0.0001 |
| *Rothia* | < 0.0001 | 4.5039 | < 0.0001 | 3.9279 | < 0.0001 |
| *Peptoniphilus* | < 0.0001 | -4.2857 | < 0.0001 | -4.4258 | < 0.0001 |
| *Lancefieldella* | < 0.0001 | 4.4368 | < 0.0001 | 3.9949 | < 0.0001 |
| *Corynebacterium* | < 0.0001 | -4.0153 | < 0.0001 | -4.5015 | < 0.0001 |
| *Fusobacterium* | < 0.0001 | 5.1968 | < 0.0001 | 4.7201 | < 0.0001 |
| *Schaalia* | < 0.0001 | 4.0294 | < 0.0001 | 4.0122 | < 0.0001 |
| *Prevotella* | < 0.0001 | 4.5406 | < 0.0001 | 4.5315 | < 0.0001 |
| *Oribacterium* | < 0.0001 | 3.9784 | < 0.0001 | 3.9614 | < 0.0001 |
| *Actinomyces* | < 0.0001 | 3.7692 | < 0.0001 | 3.6398 | < 0.0001 |
| *Megasphaera* | < 0.0001 | 3.8443 | < 0.0001 | 4.3728 | < 0.0001 |
| *Lawsonella* | < 0.0001 | -3.0156 | < 0.0001 | -3.0780 | < 0.0001 |
| *Granulicatella* | < 0.0001 | 3.8324 | < 0.0001 | 3.3532 | < 0.0001 |
| *Lachnoanaerobaculum* | < 0.0001 | 3.6079 | < 0.0001 | 3.4303 | < 0.0001 |
| *Stomatobaculum* | < 0.0001 | 3.5059 | < 0.0001 | 3.7156 | < 0.0001 |
| *Amniculibacterium* | < 0.0001 | 3.7083 | < 0.0001 | 3.2563 | < 0.0001 |
| *Centipeda* | < 0.0001 | 2.8362 | < 0.0001 | 3.3772 | < 0.0001 |
| *Streptococcus* | < 0.0001 | 2.3069 | < 0.0001 | 2.5126 | < 0.0001 |
| *Selenomonas* | < 0.0001 | 3.2467 | < 0.0001 | 3.3061 | < 0.0001 |
| *Capnocytophaga* | < 0.0001 | 4.1671 | < 0.0001 | 3.6916 | < 0.0001 |
| *Campylobacter* | < 0.0001 | 3.7907 | < 0.0001 | 3.0358 | < 0.0001 |
| *Gemella* | < 0.0001 | 3.5339 | < 0.0001 | 3.5981 | < 0.0001 |
| *Leptotrichia* | < 0.0001 | 4.2734 | < 0.0001 | 4.3430 | < 0.0001 |
| *Solobacterium* | < 0.0001 | 2.7158 | < 0.0001 | 3.0627 | < 0.0001 |
| *Treponema* | < 0.0001 | 3.7198 | < 0.0001 | 3.1574 | < 0.0001 |
| *Mogibacterium* | < 0.0001 | 2.6327 | < 0.0001 | 2.3581 | < 0.0001 |
| *Alloprevotella* | < 0.0001 | 3.4141 | < 0.0001 | 3.6683 | < 0.0001 |
| *Dolosigranulum* | < 0.0001 | -3.6113 | < 0.0001 | -3.3328 | < 0.0001 |
| *Tannerella* | < 0.0001 | 3.0864 | < 0.0001 | 2.7657 | < 0.0001 |
| *Catonella* | < 0.0001 | 2.3299 | < 0.0001 | 2.1427 | < 0.0001 |
| *Acinetobacter* | < 0.0001 | -1.8554 | < 0.0001 | -1.9548 | < 0.0001 |
| *Negativicoccus* | < 0.0001 | -1.5037 | < 0.0001 | -1.5021 | < 0.0001 |
| *Parvimonas* | < 0.0001 | 2.4248 | < 0.0001 | 2.2027 | < 0.0001 |
| *Micrococcus* | < 0.0001 | -1.2325 | < 0.0001 | -1.2240 | < 0.0001 |
| *Haemophilus* | < 0.0001 | 3.5521 | < 0.0001 | 3.6725 | < 0.0001 |
| *Dermabacter* | < 0.0001 | -0.9319 | < 0.0001 | -0.9319 | < 0.0001 |
| *Blautia* | < 0.0001 | -0.7908 | < 0.0001 | -0.7908 | < 0.0001 |
| *Eubacterium* | < 0.0001 | 2.2350 | < 0.0001 | 2.1186 | < 0.0001 |
| *Sphingomonas* | < 0.0001 | -1.1290 | < 0.0001 | -1.1176 | < 0.0001 |
| *Anaeroglobus* | < 0.0001 | 2.0345 | < 0.0001 | 1.9096 | < 0.0001 |
| *Neisseria* | < 0.0001 | 3.3592 | < 0.0001 | 3.4632 | < 0.0001 |
| *Fenollaria* | < 0.0001 | -1.0131 | < 0.0001 | -1.0221 | < 0.0001 |
| *Schwartzia* | < 0.0001 | 1.8449 | < 0.0001 | 1.3373 | < 0.0001 |
| *Kingella* | < 0.0001 | 2.1467 | < 0.0001 | 1.6429 | < 0.0001 |
| *Metamycoplasma* | < 0.0001 | 2.0195 | < 0.0001 | 1.8300 | < 0.0001 |
| *Massilia* | < 0.0001 | -0.7624 | < 0.0001 | -0.7225 | < 0.0001 |
| *Bifidobacterium* | < 0.0001 | 2.1779 | < 0.0001 | 1.6199 | < 0.0001 |
| *Methylobacterium* | < 0.0001 | -0.5405 | < 0.0001 | -0.5424 | < 0.0001 |
| *Limosilactobacillus* | < 0.0001 | 2.6458 | < 0.0001 | 1.9511 | < 0.0001 |
| *Enhydrobacter* | < 0.0001 | -0.7817 | < 0.0001 | -0.8761 | < 0.0001 |
| *Fretibacterium* | < 0.0001 | 1.8384 | < 0.0001 | 1.6790 | < 0.0001 |
| *Paracoccus* | < 0.0001 | -0.6109 | < 0.0001 | -0.5895 | < 0.0001 |
| *Kocuria* | < 0.0001 | -0.5557 | < 0.0001 | -0.5597 | < 0.0001 |
| *Porphyromonas* | < 0.0001 | 2.5838 | < 0.0001 | 2.6283 | < 0.0001 |
| *Slackia* | < 0.0001 | 1.0550 | < 0.0001 | 0.9015 | < 0.0001 |
| *Pseudomonas* | < 0.0001 | -1.3022 | < 0.0001 | -1.2923 | < 0.0001 |
| *Alloscardovia* | < 0.0001 | 1.4730 | < 0.0001 | 1.2458 | < 0.0001 |
| *Lactobacillus* | < 0.0001 | 2.5293 | < 0.0001 | 1.3835 | 0.0209 |
| *Dialister* | < 0.0001 | 1.7503 | < 0.0001 | 1.6972 | < 0.0001 |
| *Eikenella* | < 0.0001 | 1.1496 | < 0.0001 | 1.2702 | < 0.0001 |
| *Howardella* | < 0.0001 | 0.7271 | < 0.0001 | 0.7207 | < 0.0001 |
| *Shuttleworthia* | < 0.0001 | 1.0173 | < 0.0001 | 0.4902 | 0.1210 |
| *Chryseobacterium* | < 0.0001 | -0.7493 | < 0.0001 | -0.7985 | < 0.0001 |
| *Olsenella* | < 0.0001 | 1.2521 | < 0.0001 | 0.6715 | 0.0883 |
| *Pseudoramibacter* | < 0.0001 | 0.7475 | < 0.0001 | 0.1861 | 1 |
| *Arachnia* | < 0.0001 | 0.6677 | < 0.0001 | 0.4359 | 0.0209 |
| *Scardovia* | < 0.0001 | 1.3662 | < 0.0001 | 0.4970 | 0.7782 |
| *Cardiobacterium* | < 0.0001 | 1.3636 | < 0.0001 | 1.2297 | 0.0010 |
| *Brevundimonas* | < 0.0001 | -0.6708 | 0.0001 | -0.6732 | 0.0003 |
| *Cryptobacterium* | 0.0001 | 0.8959 | < 0.0001 | 0.5636 | 0.0513 |
| *Moraxella* | 0.0001 | -1.7801 | < 0.0001 | -1.2370 | 0.0244 |
| *Fudania* | 0.0003 | 0.5915 | 0.0001 | 0.1550 | 1 |
| *Peptostreptococcus* | 0.0025 | 0.6796 | 0.0345 | 0.9676 | 0.0011 |
| *Bulleidia* | 0.0062 | 0.5643 | 0.0016 | 0.3946 | 0.1309 |
| *Lautropia* | 0.0068 | 1.2377 | 0.0044 | 1.1196 | 0.0292 |
| *Moryella* | 0.0077 | 0.9238 | 0.0156 | 1.0273 | 0.0108 |
| *Butyrivibrio* | 0.0143 | 0.3580 | 0.1402 | 0.6406 | 0.0055 |
| *Filifactor* | 0.0324 | 0.7377 | 0.0139 | 0.5653 | 0.2104 |
| *Actinobacillus* | 0.0400 | 0.7870 | 0.0613 | 1.0409 | 0.0280 |

a. P values reported in the ANOVA table provide information on the term—Site’s overall effect on the model, adjusted by Holm method
b. P values represent a comparison between that specific level—(oral wash) and the reference level—(nasal), adjusted by Holm method
c. P values represent a comparison between that specific level—(sputum) and the reference level—(nasal), adjusted by Holm method

**Table E3. Taxa associated with clinical factors.**

| Clinical Factor | Corresponding Genus | Coefficient | Adjusted p value |
| --- | --- | --- | --- |
| Age | *Mannheimia* | 0.078 | 0.0014 |
| FE Phenotype | *Mogibacterium* | -0.95 | 0.029 |
| FEV1pp | *Leuconostoc* | -0.015 | 0.020 |
| Current tobacco use | *Bulleidia* | 0.49 | 0.013 |
| Pack-years of tobacco use | *Pseudomonas* | 0.016 | <0.0001 |
